# Systemic hypoxemia catalyzes cerebral oxidative-nitrosative stress during extreme apnea in humans: implications for cerebral bioenergetic function

**DOI:** 10.1101/2023.07.24.23293124

**Authors:** Damian M. Bailey, Anthony R. Bain, Ryan L. Hoiland, Otto F. Barak, Ivan Drvis, Benjamin S. Stacey, Angelo Iannetelli, Gareth W. Davison, Rasmus H. Dahl, Ronan M.G. Berg, David B. MacLeod, Zeljko Dujic, Philip N. Ainslie

## Abstract

**BACKGROUND:** Voluntary asphyxia induced by apnea in competitive breath hold (BH) divers affords a unique opportunity to examine integrated mechanisms underlying the preservation of cerebral bioenergetic function. This study examined to what extent physiological extremes of oxygen (O_2_) demand and carbon dioxide (CO_2_) production impact redox homeostasis and corresponding red blood cell (RBC)-mediated cerebral vasodilation.

**METHODS:** Ten ultra-elite apneists (6 men, 4 women) aged 33 ± 9 (mean ± SD) years old performed two maximal dry apneas preceded by, [1] normoxic normoventilation resulting in severe hypoxemic hypercapnia apnea (HHA) and [2] hyperoxic hyperventilation designed to prevent hypoxemia resulting in isolated hypercapnic apnea (IHA). Transcerebral exchange kinetics of ascorbate radicals (A^·-^, electron paramagnetic resonance spectroscopy), lipid hydroperoxides (LOOH, spectrophotometry) and nitric oxide metabolites (NO, tri-iodide reductive chemiluminescence) were calculated as the product of global cerebral blood flow (gCBF, duplex ultrasound) and radial arterial (a) to internal jugular venous (v) concentration gradients determined at eupnea and after apnea.

**RESULTS:** Apnea duration increased from 306 ± 62 s during HHA to 959 ± 201 s during IHA (P = <0.001), resulting in individual nadirs of 29 mmHg and 40 % for PaO_2_ and SaO_2_ respectively in HHA and PaCO_2_ peak of 68 mmHg in IHA. Apnea resulted in a more pronounced elevation in the net cerebral output (v>a) of A^·-^ and LOOH in HHA (P = <0.05 vs. IHA). This coincided with a lower apnea-induced increase in gCBF (P = <0.001 vs. IHA) and related suppression in plasma nitrite (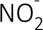) uptake (a>v) (P = < 0.05 vs. IHA), implying reduced consumption and delivery of NO consistent with elevated cerebral oxidative-nitrosative stress (OXNOS). While apnea-induced gradients consistently reflected plasma 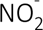 consumption (a>v) and RBC iron nitrosylhemoglobin formation (v>a), we failed to observe equidirectional gradients consistent with *S*-nitrosohemoglobin consumption and plasma *S*-nitrosothiol delivery.

**CONCLUSIONS:** These findings highlight a key catalytic role for hypoxemia in cerebral OXNOS with 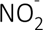 reduction the more likely mechanism underlying endocrine NO vasoregulation with the capacity to transduce physiological O_2_-CO_2_ gradients into graded vasodilation.

## INTRODUCTION

The neurovascular complex (NVC) is a recent concept that highlights the functional interaction between the multi-hetero-cellular structure comprizing the neurovascular unit (NVU) and the systemic vasculature^1^. Precisely how the NVC integrates, coordinates and processes molecular signals that collectively maintain cerebral blood flow (CBF) and structural integrity of the blood–brain barrier (BBB) remains to be fully understood, although emergent evidence suggests a key role for redox-regulation^2^.

In support, a neuroprotective hormetic role has emerged for the physiological formation of free radicals and associated reactive oxygen/nitrogen species (ROS/RNS) that enable the sensing and subsequent clearance/delivery of the respiratory gases carbon dioxide (CO_2_) and oxygen (O_2_) to which the human brain has evolved exquisite sensitivity. In contrast, when excessive and sustained, free radical formation can result in structural destabilization and vascular impairment of the NVU subsequent to oxidative inactivation of vascular nitric oxide (NO), collectively termed oxidative-nitrosative stress (OXNOS)^3, 4^.

However, to what extent O_2_ and CO_2_ tensions independently modulate NVC function including the potentially differential impact on the redox-regulation of nitrite (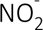) and *S-* nitrosohemoglobin (SNO-Hb) bioactivity, the principal competing mechanisms underlying endocrine NO vasoregulation^5^, remains to be established. Prolonged apnea performed by ultra-elite breath-hold (BH) divers, given its ability to evoke profound hypoxemic-hypercapnia and compensatory adaptations that preserve bioenergetic homeostasis, affords a unique opportunity to address these knowledge gaps.

Extending prior work focused exclusively on BBB integrity^6^, this study combined volumetric assessment of CBF with concurrent sampling of arterio-jugular venous concentration differences (a-v_D_) to calculate cerebral exchange kinetics of intravascular OXNOS biomarkers in world-class apneists over the course of two maximal, static apneas performed in ambient air. The first was performed following (standard) prior normoxic normoventilation resulting in severe hypoxemia-hypercapnia apnea (HHA) whereas the second followed prior hyperoxic hyperventilation, designed to prevent hypoxemia resulting in isolated hypercapnic apnea (IHA). By manipulating arterial O_2_ independent of CO_2_ tension, we sought to address the following hypotheses (Figure 1A). First, that cerebral output [venous (v) > arterial (a)] of free radicals and suppression of NO metabolite consumption and CBF would be more pronounced in the normoxic (HHA) compared to hyperoxic (IHA) apnea, implicating hypoxemia as the dominant OXNOS catalyst. Second that the apnea-induced elevation in CBF would be proportional to a net loss (a > v) of 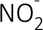 and SNO-Hb taken to reflect metabolite consumption and delivery of NO.

**Figure 1.**
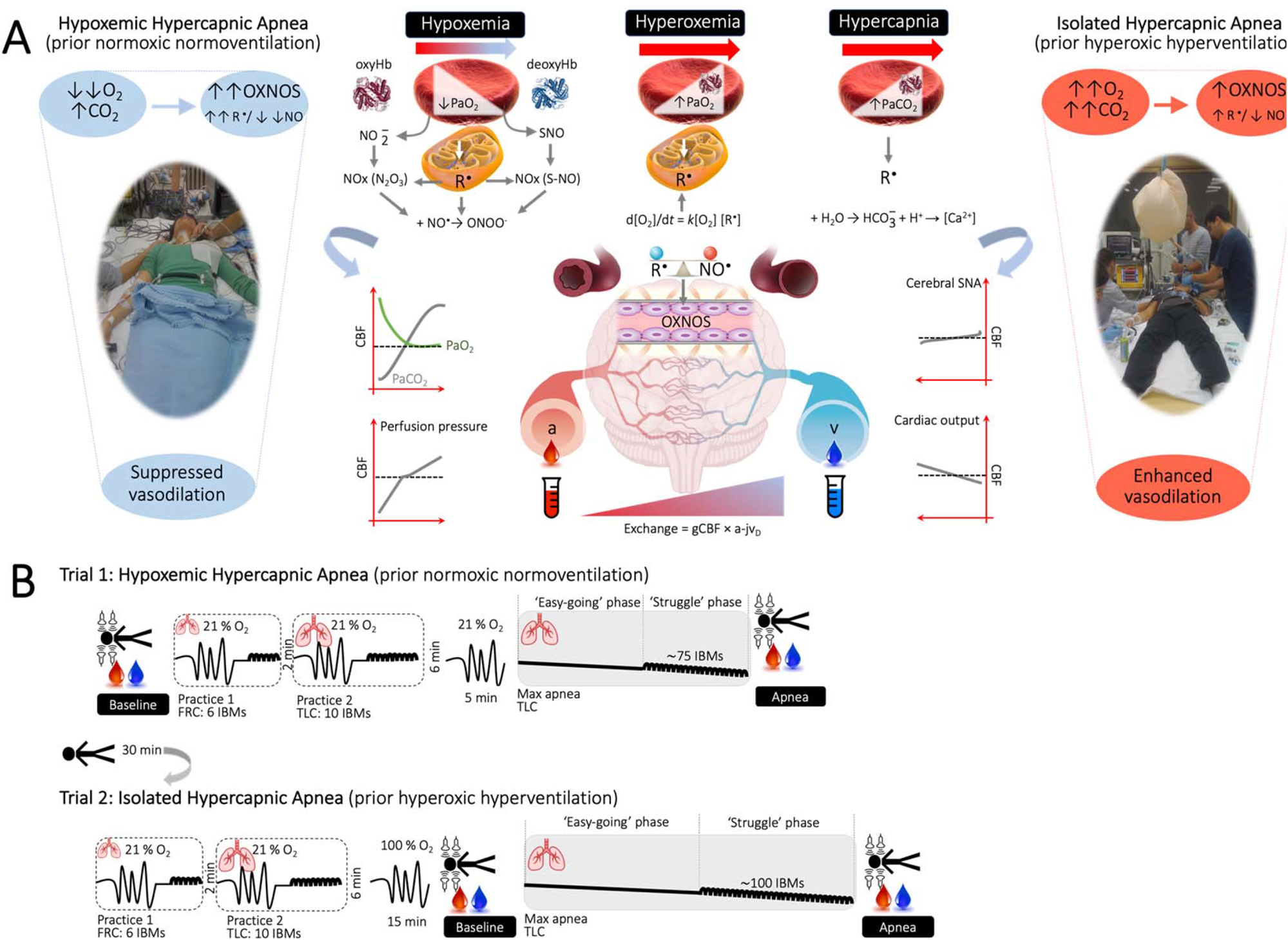
Experimental hypotheses (A) and design (B) A. Predicted differential disruption of cerebral redox homeostasis and implications for cerebral bioenergetic function in response to pathological extremes of oxygen (O_2_) and carbon dioxide (CO_2_) during hypoxemic hypercapnia (HHA, prior normoxic normoventilation) and isolated hyperoxemic hypercapnia (IHA, prior hyperoxic hyperventilation) apneas. O_2_/CO_2_ sensing by the red blood cell (RBC) and mitochondrion is allosterically coupled to formation of nitric oxide (NO^•^)^43^ and reactive oxygen/nitrogen (ROS/RNS)^44^ species that titrate (global) cerebral blood flow (gCBF) coupling substrate (O_2_ and glucose) delivery to demand. The more marked reduction in arterial partial pressure of oxygen (PO_2_) during the HHA trial is hypothesized to induce a more pronounced elevation in cerebral oxidative-nitrosative stress (OXNOS) reflected by a free radical-mediated reduction in vascular NO bioavailability (manifest as increased net cerebral output of free radicals and lipid peroxidants and corresponding lower net cerebral uptake of nitrite (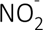) or *S*-nitrosohemoglobin (SNO-Hb) leading to a suppression of cerebral hypoxic vasodilation relative to IHA trial. Note typical human experimental setup highlighting arterial-jugular venous (transcerebral) sampling of blood combined with volumetric assessment of gCBF. R^•^, free radical; 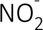, nitrite; N_2_O_3_, nitrogen tri-oxide; ONOO^-^, peroxynitrite (not measured); PaO_2_, arterial partial pressure of oxygen; PaCO_2_, arterial partial pressure of carbon dioxide; a-jv_D_, arterio-jugular venous concentration difference; SNA, sympathetic nervous activity (not measured). B. Two submaximal (practice) apneas preceded separate trials involving prior normoxic (21 % O_2_) normoventilation resulting in severe hypoxemic hypercapnia apnea (HHA) whereas the second followed prior hyperoxic hyperventilation (100 % O_2_), designed to prevent hypoxemia resulting in isolated hypercapnic apnea (IHA). The first practice apnea was performed at functional residual capacity (FRC) for 6 involuntary body movements (IBMs) and the second practice at total lung capacity (TLC) for 10 IBMs. Both maximal apneas were performed at TLC. Measurements were performed at baseline and timed to coincide with the end of each maximal apnea.

## METHODS

### Ethics

Experimental procedures were approved by the University of Split, Ethics Committee (#H14-00922). All procedures were carried out in accordance with the most (7^th^) recent amendment of the Declaration of Helsinki of the World Medical Association^7^ (with the exception that it was not registered in a publicly accessible database prior to recruitment) with verbal and written informed consent obtained from all participants.

### Participants

Ten physically active, elite apneists (6 men, 4 women) aged 33 (mean) ± 9 (SD) years old with a body mass index of 23 ± 2 kg/m^2^ and forced vital capacity of 6.40 ± 1.48 L volunteered from the Croatian National Apnea Team. At the time of these experiments, six had placed within the world’s top 10 within the last 5 years. One female set a new official world record in dynamic apnea while another male set the world record for apnea following prior hyperoxic hyperventilation (24 min 33 s). A medical examination confirmed that all participants were free of cardiovascular, pulmonary and cerebrovascular disease and were not taking any nutritional supplements including over-the-counter antioxidant or anti-inflammatory medications. They were advised to refrain from physical activity, caffeine and alcohol for 24 h prior to formal experimentation and maintain their normal dietary behavior. Participants attended the laboratory following a 12 h overnight fast.

### Design

Parts of this study including blood gas and select aspects of cerebral hemodynamic function (Supplementary material Tables 1-2) have previously been published as part of separate investigations focused on the link between hypercapnia and cerebral oxidative metabolism^8^ and hyper-perfusion-mediated structural destabilization of the NVU^6^. Thus, although the present study adopted an identical experimental design, it constitutes an entirely separate investigation complemented by *de novo* experimental measures and *a priori* hypotheses focused exclusively on altered redox homeostasis. Participants were required to perform two maximal apneas preceded by, [1] normoxic normoventilation resulting in severe hypoxemic hypercapnia apnea (HHA) and [2] hyperoxic hyperventilation designed to prevent hypoxemia resulting in isolated hypercapnic apnea (IHA). The order of trials was non-randomized given the long-lasting carryover effects of hyperoxia and fatigue associated with a more prolonged apnea^8^. Data were collected at baseline and timed to coincide with the point of apnea termination (Figure 1B).

### Procedures

All apneas were completed on a single day with the Croatian national apnea coach present to motivate divers and ensure maximal efforts. Each apnea was preceded by 30 min of supine rest followed by two standardized preparatory (practice) apneas designed to maximize the experimental apneas (see below). The first preparatory apnea was performed at functional residual capacity (FRC) until seven involuntary breathing movements (IBMs) were attained. Two minutes later, the second preparatory apnea was performed at total lung capacity (TLC) lasting for ten IBMs. Participants then rested quietly for 6 min prior to two ‘maximal’ apneas that were each performed at TLC:

Trial 1: Prior normoxic normoventilation-hypoxemic hypercapnia apnea (HHA)

Participants performed a maximal apnea in room air (normoxia).

Trial 2: Prior hyperoxic hyperventilation-isolated hypercapnic apnea (IHA)

Participants performed a maximal apnea preceded by 15 min of hyperventilation of 100% O_2_ while receiving auditory feedback from the coach to achieve an end tidal PCO_2_ (PET_CO2_) of ∼20 mmHg.

### Blood sampling

#### Catheterization

Participants were placed slightly head down on a hospital bed, and two catheters were inserted retrograde using the Seldinger technique under local anesthesia (1 % lidocaine) via ultrasound guidance. A 20-guage arterial catheter (Arrow, Markham, ON, Canada) was placed in the right radial artery (a) and attached to an in-line waste-less sampling setup (Edwards Lifesciences VAMP, CA, USA) connected to a pressure transducer that was placed at the height of the right atrium (TruWave transducer). A central venous catheter (Edwards PediaSat Oximetry Catheter, CA, USA) was placed in the right internal jugular vein (v) and advanced towards the jugular bulb located at the mastoid process, approximately at the C1-C2 interspace (Figure 1). Facial vein contamination was ruled out by ensuring that all jugular venous oxyhemoglobin saturation (SO_2_) values were < 75% at rest. Participants rested for at least 30 min following catheter placement prior to the collection of baseline samples.

#### Collection and storage

Blood samples were drawn without stasis simultaneously from the RA and JV directly into Vacutainers (Becton, Dickinson and Company, Oxford, UK) before centrifugation at 600g (4°C) for 10 min. With the exception of blood for determination of blood gas variables, plasma and red blood cell (RBC) samples were decanted into cryogenic vials (Nalgene Labware, Thermo Fisher Scientific Inc, Waltham, MA) and immediately snap-frozen in liquid nitrogen (N_2_) before transport under nitrogen gas (Cryopak, Taylor-Wharton, Theodore, AL) from Croatia to the United Kingdom. Samples were left to defrost at 37°C in the dark for 5 minutes prior to batch analysis.

### Measurements

#### Blood gases

Whole blood was collected into heparinized syringes, maintained anerobically at room temperature and immediately analyzed for hemoglobin (Hb), hematocrit (Hct), partial pressures of oxygen, carbon dioxide (PO_2_/PCO_2_), SO_2_, pH and glucose (Glu) using a commercially available cassette-based analyzer (ABL-90 FLEX, Radiometer, Copenhagen, Denmark). Hydrogen ion (H^+^) concentration was calculated as [H^+^] = 10^-pH^. The intra- and inter-assay CVs for all metabolites were both <5%^9^.

#### Antioxidants

Plasma was stabilized and deproteinated using 10% metaphosphoric acid (Sigma Chemical, Dorset, UK). Ascorbic acid was assayed by fluorimetry based on the condensation of dehydroascorbic acid with 1,2-phenylenediamine^10^. Concentrations of the lipid soluble antioxidants (LSA) encompassing α/γ-tocopherol, α/β-carotene, retinol and lycopene were determined using high-performance liquid chromatography (HPLC)^11, 12^. The intra- and inter-assay CVs for both metabolites were both <5%^9^.

#### Free radicals

*Ascorbate free radical (A^·-^)*: We employed electron paramagnetic resonance (EPR) spectrosopic detection of A^·-^ as a direct measure of global systemic free radical formation^9^. Plasma (1 mL) was injected directly into a high-sensitivity multiple-bore sample cell (AquaX, Bruker Daltonics Inc., Billerica, MA, USA) housed within a TM_110_ cavity of an EPR spectrometer operating at X-band frequency (9.87 GHz). Samples were recorded by cumulative signal averaging of 10 scans using the following instrument parameters: resolution, 1024 points: microwave power, 20 mW; modulation amplitude, 0.65 G; receiver gain, 2 × 10^6^; time constant, 40.96 ms; sweep rate, 0.14 G/s; sweep width, 6 G; centre field, 3486 G. All spectra were filtered identically (moving average, 15 conversion points) using WINEPR software (Version 2.11, Bruker, Karlsruhe, Germany) and the double integral of each doublet quantified using Origin 8 software (OriginLab Corps, MA, USA). The intra- and inter-assay CVs were both <5%^9^.

*Lipid hydroperoxides (LOOH)*: LOOH were determined spectrophotometrically via the ferric oxidation of xylenol orange (FOX) version II method, with modification^9^. Intra- and inter-assay CVs were <2 % and <4 % respectively^9^.

#### Nitric oxide (NO) metabolites

Ozone-based chemiluminescence (Sievers NOA 280i, Analytix Ltd, Durham, UK) was used to detect NO liberated from plasma and RBC samples via chemical reagent cleavage facilitating detection of plasma and red blood cell (RBC) 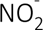, *S*-nitrosothiols (RSNO), *S*-nitrosohemoglobin (SNO-Hb) and iron nitrosylhemoglobin (HbNO) according to established methods^13–15^. Total NO bioactivity was calculated as the cumulative concentration of plasma and RBC metabolites. Signal output was plotted against time using Origin 8 software (OriginLab Corps, Massachusetts, USA) and smoothed using a 150-point averaging algorithm. The Peak Analysis package was used to calculate the area under the curve and subsequently converted to a concentration using standard curves of sodium 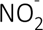. Intra- and inter-assay CVs for all metabolites were both <10%^13^.

#### Cardiopulmonary function

All cardiopulmonary measurements were averaged over 15 s immediately prior to blood sampling. A lead II electrocardiogram (Dual BioAmp; ADInstruments, Oxford, UK) was used to measure heart rate (HR). Intra-arterial ABP was recorded directly via a pressure transducer placed at the level of the heart for determination of mean arterial pressure (MAP). Internal jugular venous pressure (IJVP) was measured with the transducer connected to the jugular catheter. End-tidal partial pressures of oxygen and carbon dioxide (PET_O2/CO2_) were measured via capnography (ML 206, ADInstruments Ltd, Oxford, UK). Finger photoplethysmography (Finometer PRO, Finapres Medical Systems, Amsterdam, The Netherlands) was used to measure beat-by-beat stroke volume (SV) and cardiac output (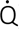) using the Modelflow algorithm^16^ that incorporates participant sex, age, stature and mass (BeatScope 1.0 software; TNO; TPD Biomedical Instrumentation, Amsterdam, The Netherlands).

#### Cerebrovascular function

*Intracranial:* Blood velocity in the middle cerebral artery (MCAv, insonated through the left temporal window and posterior cerebral artery (PCAv, insonated at the P1 segment through the right temporal window) were measured using standardized procedures with a 2 MHz pulsed transcranial Doppler ultrasound (TCD; Spencer Technologies, Seattle, WA, USA). Bilateral TCD probes were secured using a specialized commercial headband (Mark600, Spencer Technologies, Seattle, WA, USA) using standardized search techniques.

*Extracranial:* Continuous diameter, velocity and blood flow recordings in the right internal carotid and left vertebral arteries 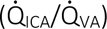 were obtained using a 10 MHz, multifrequency, linear array vascular ultrasound (Terason 3200, Teratech, Burlington, MA). Arterial diameter was measured via B-mode imaging, whereas peak blood velocity was simultaneously measured with pulse-wave mode. The ICA was insonated ≥1.5 cm from the carotid bifurcation, with no evidence of turbulent or retrograde flow present during recording. The VA was insonated at the C4–C5 or C5–C6 vertebral segment and standardized within participants for the HHA and IHA trials. The steering angle was fixed to 60° and the sample volume was placed in the center of the vessel and adjusted to cover the entire vascular lumen. All images were recorded as video files at 30 Hz and stored for offline analysis using customized edge detection software designed to mitigate observer bias^17^. Simultaneous measures of arterial diameter and velocity over >12 consecutive cardiac cycles were used to calculate flow. Between-day CVs for 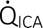 and 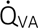 are 5 % and 11 %, respectively^18^.

### Data integration

All cardiopulmonary measurements were sampled at 1 kHz and integrated into PowerLab® and LabChart® software (ADInstruments, Bella Vista, NSW, Australia) for online monitoring and saved for offline analysis.

### Calculations

#### Cerebral bioenergetics

*Perfusion:* Volumetric blood flow was calculated offline as:

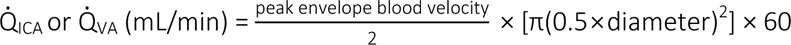

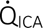 or 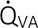 were averaged for a 1 min eupnea baseline and during the last 30 s of apnea.

During the latter half of each apnea, IBMs and corresponding contraction of the sternocleidomastoid prevented reliable blood velocity traces. To circumvent this, Q_ICA_ and Q_VA_ were estimated (e) from changes in MCAv (for 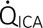) and PCAv (for 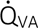) respectively, as previously described^19, 20^:

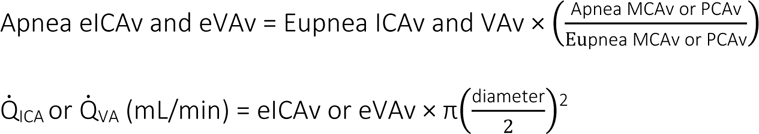

Assuming symmetrical blood flow of contralateral ICA and VA arteries, global cerebral blood flow (gCBF) was calculated as:

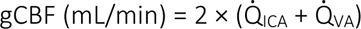

Cerebral perfusion pressure (CPP) was calculated as MAP-IJVP with the latter serving as a surrogate for ICP^21^.

*Metabolism:* Arterial and jugular venous oxygen content (caO_2_ and cvO_2_ respectively) were calculated as:

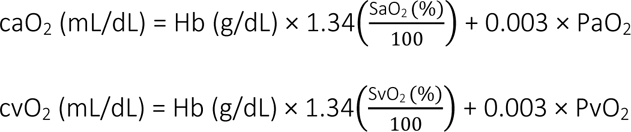

where 1.34 is the O_2_ binding capacity of Hb and 0.003 is the solubility of O_2_ dissolved in blood.

Global cerebral O_2_ and glucose delivery (gCDO_2_/gCD_Glucose_) were calculated as:

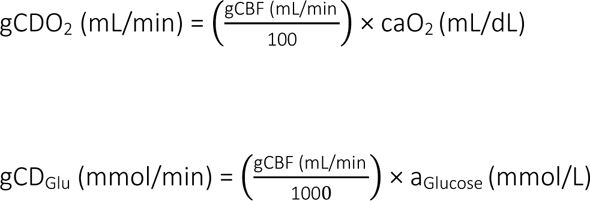

The cerebral metabolic rate of oxygen (CMRO_2_) and glucose (CMR_Glucose_) were calculated as:

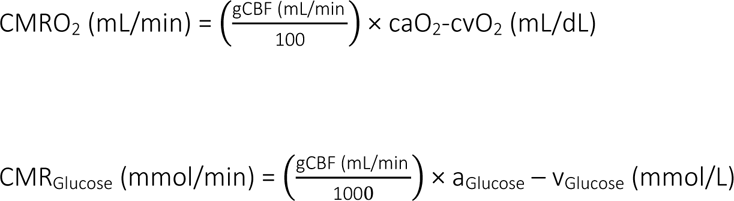

#### Transcerebral exchange kinetics

gCBF was converted into global cerebral plasma (gCPF) and red blood cell (gCRBCF) flow:

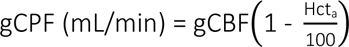

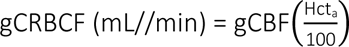

for corresponding calculation of net transcerebral exchange kinetics of plasma/serum/RBC-borne metabolites according to the Fick principle:
Exchange (µmol or nmol/min) = gCBF or gCPF or gCRBCF × (a – v_D_).

By convention, a positive value (a>v) refers to net uptake (loss or consumption) whereas a negative value (v>a) indicates net output (gain or formation) across the cerebrovascular bed.

### Data analysis

Data were analyzed using the Statistics Package for Social Scientists (IBM SPSS Statistics Version 29.0). Distribution normality was confirmed using Shapiro-Wilk *W* tests (*P* > 0.05). Data were analyzed using a combination of two (*State*: Eupnea *vs*. Apnea × *Site*: Arterial *vs*. Venous) and three (*Trial*: HHA *vs*. IHA × *State*: Eupnea *vs*. Apnea × *Site*: Arterial *vs*. Venous) factor repeated measures analyzes of variance. Post-hoc Bonferroni-corrected paired samples *t*-tests were employed to locate differences following an interaction. Relationships between selected variables were analyzed using Pearson Product Moment Correlations. Significance was established at *P* < 0.05 for all two-tailed tests and data presented as mean ± SD.

## RESULTS

### Apnea

Apnea duration increased from 306 ± 62 s (range: 217-409 s) during HHA to 959 ± 201 s (range: 579-1262 s) during IHA (P = <0.001).

### Blood gas variables

As expected, apnea induced severe systemic and local hypoxemia (↓PO_2_, ↓SO_2_, ↓cO_2_), hypercapnia (↑PCO) and acidosis 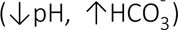 in NX (Supplemental Table 1). In one participant, nadirs of 29 mmHg and 40 % were recorded for PaO_2_ and SaO_2_ respectively, corresponding to a (peak) PaCO_2_ of 53 mmHg in HHA. Participants were hypocapnic at eupnea in IHA and remained hyperoxemic and comparatively less hypertensive and more hypercapnic/acidotic throughout apnea (Supplemental Table 1). The highest individual value recorded for PaCO_2_ during IHA was 68 mmHg corresponding to a pH of 7.227. Baseline levels of Hb and Hct at eupnea were elevated in IHA (Supplemental Table 1).

### Cardiopulmonary function

Apnea decreased SV and 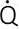 in HHA whereas increases were observed in IHA (Table 1). Apnea was associated with a general increase in MAP, IJVP and CPP that were more pronounced in HHA (Supplemental Table 1).

**Table 1.** Oxidative stress. Values are mean ± SD; a-v_D_, arterio-jugular venous concentration difference; Total lipid soluble antioxidants (LSA) calculated as the cumulative concentration of α/γ-tocopherol, α/β-carotene, retinol and lycopene; A^·-^, ascorbate free radical; LOOH, lipid hydroperoxides. Positive or negative a-v_D_ reflects cerebral uptake (loss or consumption) or output (gain or formation) respectively.

| Trial: | Hypoxemic Hypercapnia Apnea (n = 10) |  |  |  | Isolated Hypercapnic Apnea (n = 10) |  |  |  |
| --- | --- | --- | --- | --- | --- | --- | --- | --- |
| State: | Eupnea |  | Apnea |  | Eupnea |  | Apnea |  |
| Site: | Arterial | Venous | Arterial | Venous | Arterial | Venous | Arterial | Venous |
| Antioxidants |  |  |  |  |  |  |  |  |
| Ascorbate (μM) | 59.3 ± 11.0 | 59.2 ± 10.0 | 61.8 ± 8.9 | 53.5 ± 6.4 | 61.0 ± 6.7 | 60.5 ± 5.3 | 63.0 ± 5.6 | 60.0 ± 3.7 |
| Site (P = 0.007); Trial × Site (P = 0.017); State × Site (P = 0.015) |  |  |  |  |  |  |  |  |
| a-v<sub>D</sub> (μM) | 0.2 ± 4.3 |  | 8.3 ± 5.6 |  | 0.5 ± 4.9 |  | 3.0 ± 4.0 |  |
| Trial (P = 0.017); State (P = 0.015) |  |  |  |  |  |  |  |  |
| α-tocopherol (μM) | 22.1 ± 3.9 | 21.7 ± 3.2 | 24.4 ± 3.0 | 20.4 ± 3.4 | 22.5 ± 4.9 | 22.7 ± 4.1 | 22.7 ± 2.9 | 21.3 ± 3.2 |
| Site (P = 0.008); State × Site (P = 0.011) |  |  |  |  |  |  |  |  |
| a-v<sub>D</sub> (μM) | 0.3 ± 1.5 |  | 4.0 ± 2.6 |  | -0.1 ± 4.2 |  | 1.4 ± 2.6 |  |
| State (P = 0.010) |  |  |  |  |  |  |  |  |
| Total LSA (μM) | 25.6 ± 3.6 | 25.2 ± 3.1 | 28.0 ± 2.6 | 23.7 ± 3.0 | 26.0 ± 4.7 | 26.2 ± 3.9 | 26.0 ± 2.5 | 24.4 ± 2.7 |
| Site (P = 0.006); State × Site (P = 0.010) |  |  |  |  |  |  |  |  |
| a-v<sub>D</sub> (μM) | 0.4 ± 2.1 |  | 4.3 ± 2.4 |  | -0.1 ± 4.7 |  | 1.6 ± 2.6 |  |
| State (P = 0.011) |  |  |  |  |  |  |  |  |
| Free radicals |  |  |  |  |  |  |  |  |
| A<sup>•</sup> (AU × 10<sup>2</sup>) | 320 ± 107 | 313 ± 98 | 344 ± 95 | 473 ± 127 | 322 ± 70 | 333 ± 102 | 363 ± 143 | 386 ± 129 |
| State (P = 0.009); Site (P = 0.046); State × Site (P = 0.048) |  |  |  |  |  |  |  |  |
| a-v<sub>D</sub> (AU × 10<sup>2</sup>) | 6 ± 130 |  | -129 ± 96 |  | -12 ± 143 |  | -24 ± 91 |  |
| State (P = 0.048) |  |  |  |  |  |  |  |  |
| Lipid peroxidation |  |  |  |  |  |  |  |  |
| LOOH (μM) | 1.15 ± 0.17 | 1.16 ± 0.13 | 1.22 ± 0.25 | 1.74 ± 1.20 | 1.19 ± 0.15 | 1.25 ± 0.22 | 1.27 ± 0.18 | 1.45 ± 0.24 |
| State (P = 0.038); Site (P = 0.045); State × Site (P = 0.048) |  |  |  |  |  |  |  |  |
| a-v<sub>D</sub> (μM) | -0.01 ± 0.14 |  | -0.52 ± 0.97 |  | -0.07 ± 0.23 |  | -0.18 ± 0.15 |  |
| State (P = 0.047) |  |  |  |  |  |  |  |  |

### Cerebral bioenergetics

Compared to the eupnic control states (normoxic normocapnia), a greater increase in gCBF was observed in IHA (+206 ± 52 %) compared to HHA (+83 ± 22 %, *P* = <0.001) due to consistently lower 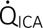 and 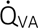 values at baseline subsequent to (hyperoxic) hyperventilation-induced hypocapnia (Supplemental Table 3). Corresponding increases in local (VA/ICA) and global cerebral substrate delivery (O_2_ and glucose) were also more pronounced during apnea in IHA (Supplemental Table 3). Apnea generally decreased CMRO_2_ whereas CMR_Glu_ remained preserved with no differences between trials (Supplemental Table 3).

### Oxidative stress

Apnea increased the a-v_D_ and corresponding net cerebral uptake of ascorbate, α-tocopherol and total LSA that were more pronounced in HHA (Table 1, Figure 2 A-C). This was accompanied by a reduction in the a-v_D_ and reciprocal elevation in the net cerebral output of A^·-^ and LOOH that were also more marked in HHA for A^·-^ (Table 1, Figure 2 D-E).

**Figure 2.**
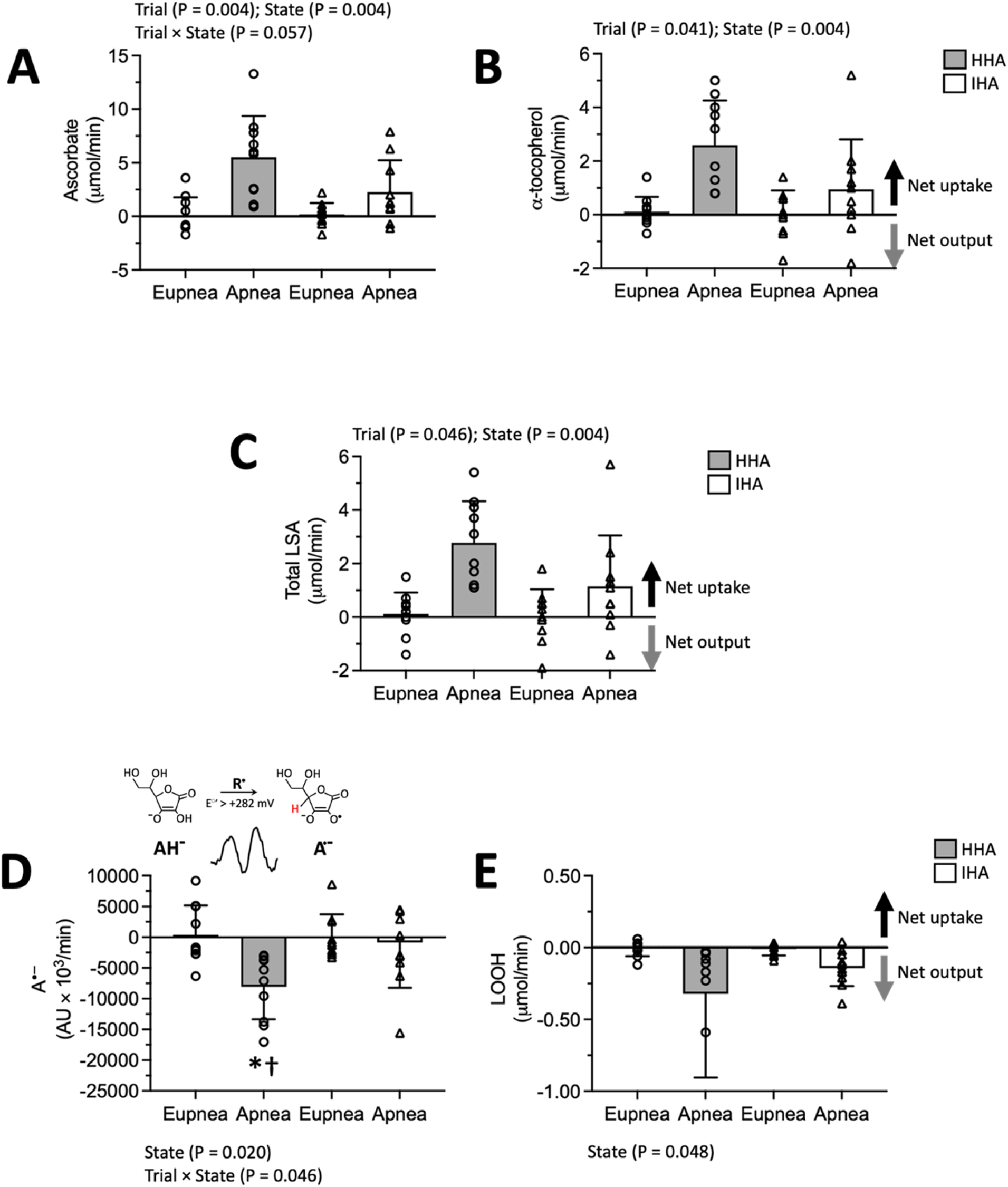
Transcerebral exchange of oxidative stress biomarkers. Values are mean ± SD; Separate trials involving prior normoxic (21 % O_2_) normoventilation resulting in severe hypoxemic hypercapnia apnea (HHA) whereas the second followed prior hyperoxic (100 % O_2_) hyperventilation, designed to prevent hypoxemia resulting in isolated hypercapnic apnea (IHA); Circles and triangles denote individual data points for participants during the NX and HX trials respectively; Total lipid soluble antioxidants (LSA) calculated as the cumulative concentration of α/γ-tocopherol, α/β-carotene, retinol and lycopene; A^·-^, ascorbate free radical; LOOH, lipid hydroperoxides. Exchange calculated as the product of global cerebral plasma/serum flow and arterio-jugular venous concentration difference. Positive or negative gradients reflect net cerebral uptake (loss or consumption) or output (gain or formation) respectively. Since the concentration of ascorbate in human plasma is orders of magnitude greater than any oxidizing free radical combined with the low one-electron reduction potential for the A^•-^/ascorbate monanion (AH^-^) couple (*E*^°^΄ = 282 mV)^45^, any oxidizing species (R^•^) generated locally within the cerebral circulation will result in the one-electron oxidation of ascorbate to form the distinctive EPR-detectable A^•-^ doublet (R^•^ + AH^-^ → A^•-^ + R-H, Panel D inset with main coupling hydrogen highlighted in red)^46^. *different between state for given trial (*P* < 0.05); †different between trial for given state (*P* < 0.05).

### Nitrosative stress

Figure 3 highlights the dynamic interplay and re-apportionment of NO metabolites across the cerebral circulation observable within the constraints of a single a-v transit. The total NO metabolite pool was generally lower in HHA and during apnea due to a combined reduction in plasma and RBC 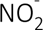 whereas elevations were observed in SNO-Hb and HbNO (Table 2, Figure 3). Corresponding differences translated into net cerebral uptake of plasma 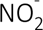, RSNO and RBC 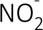 (Figure 4 A-B, D) that were opposed by net cerebral output of SNO-Hb and HbNO (Figure 4 E-F). Plasma 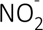 uptake and SNO-Hb/HbNO output were more pronounced during apnea, with plasma 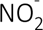uptake generally lower in HHA (Figure 4 A, E-F). The apnea-induced elevation in A^·-^ output was associated with a reduction in plasma 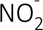 uptake in HHA (r = 0.702, P = 0.024) but not IHA (r = 0.509, P = 0.133).

**Figure 3.**
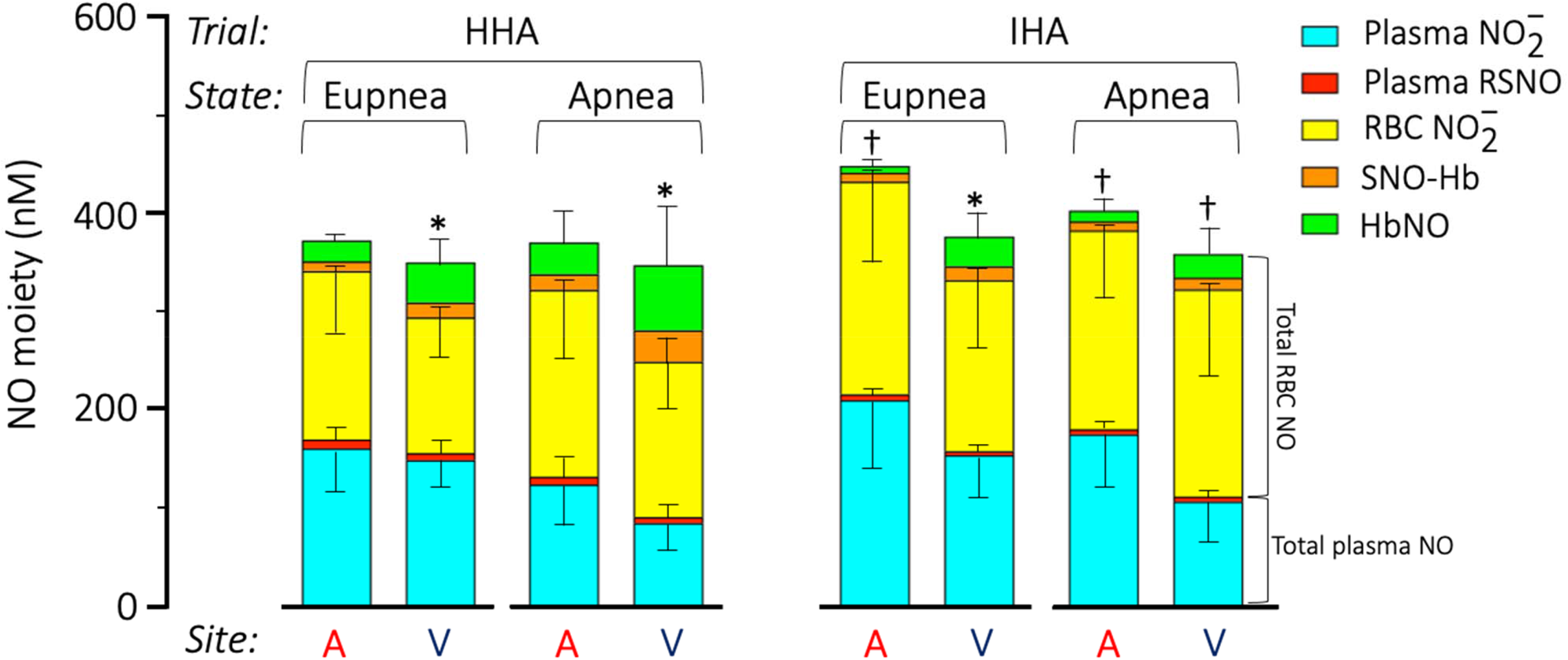
Local distribution of nitric oxide (NO) metabolites. Values are mean ± SD; Separate trials involving prior normoxic (21 % O_2_) normoventilation resulting in severe hypoxemic hypercapnia apnea (HHA) whereas the second followed prior hyperoxic hyperventilation (100 % O_2_), designed to prevent hypoxemia resulting in isolated hypercapnic apnea (IHA); NO, nitric oxide 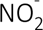, nitrite; RSNO, *S*-nitrosothiols; SNO-Hb, *S*-nitrosohemoglobin; HbNO, nitrosylhemoglobin; A, arterial; V, venous. Plasma 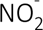: *Trial (P = 0.010); State (P = 0.003); Site (P = <0.001); Trial × Site (P = 0.048);* RBC 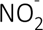: *Trial (P = 0.048); Site (P = 0.047);* SNO-Hb: *Trial (P = 0.014); State (P = 0.036); Site (P = 0.019); Trial × State (P = 0.022);* HbNO: *Trial (P = 0.002); Site (P = <0.001); Trial × State (P = 0.024); Trial × State × Site (P = 0.045);* Total plasma NO: *Trial (P = 0.009); State (P = 0.033); Site (P = <0.001); Trial × Site (P = 0.042);* Total RBC NO: *State (P = 0.042);* Total plasma + RBC NO: *Trial (P = 0.044); Site (P = 0.036).* *different between state for given trial (*P* < 0.05); †different between trial for given state (*P* < 0.05).

**Figure 4.**
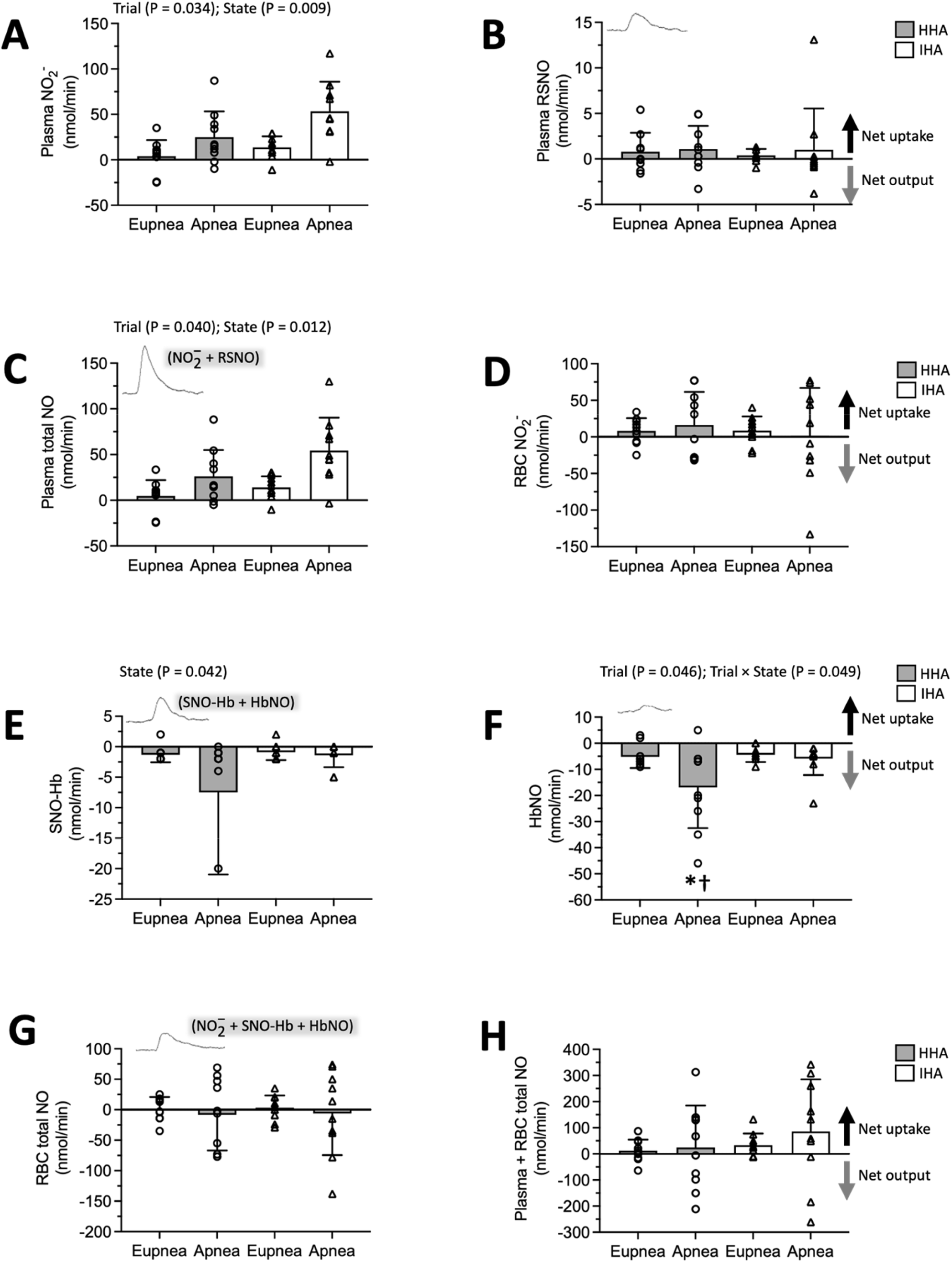
Transcerebral exchange of nitrosative stress biomarkers. Values are mean ± SD; Separate trials involving prior normoxic (21 % O_2_) normoventilation resulting in severe hypoxemic hypercapnia apnea (HHA) whereas the second followed prior hyperoxic hyperventilation (100 % O_2_), designed to prevent hypoxemia resulting in isolated hypercapnic apnea (IHA); Circles and triangles denote individual data points for participants during the NX and HX trials respectively; NO, nitric oxide; 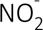, nitrite; RSNO, *S*-nitrosothiols; SNO-Hb, *S*-nitrosohemoglobin; HbNO, nitrosylhemoglobin. Exchange calculated as the product of global cerebral plasma (A-C) or red blood cell (RBC, D-G) or whole blood (H) flow and arterio-jugular venous concentration difference. Positive or negative gradients reflect net cerebral uptake (loss or consumption) or output (gain or formation) respectively. Note typical smoothed traces for arterial concentration of metabolites obtained by ozone-based chemiluminescence (same scale). *different between state for given trial (*P* < 0.05); †different between trial for given state (*P* < 0.05).

**Table 2.**
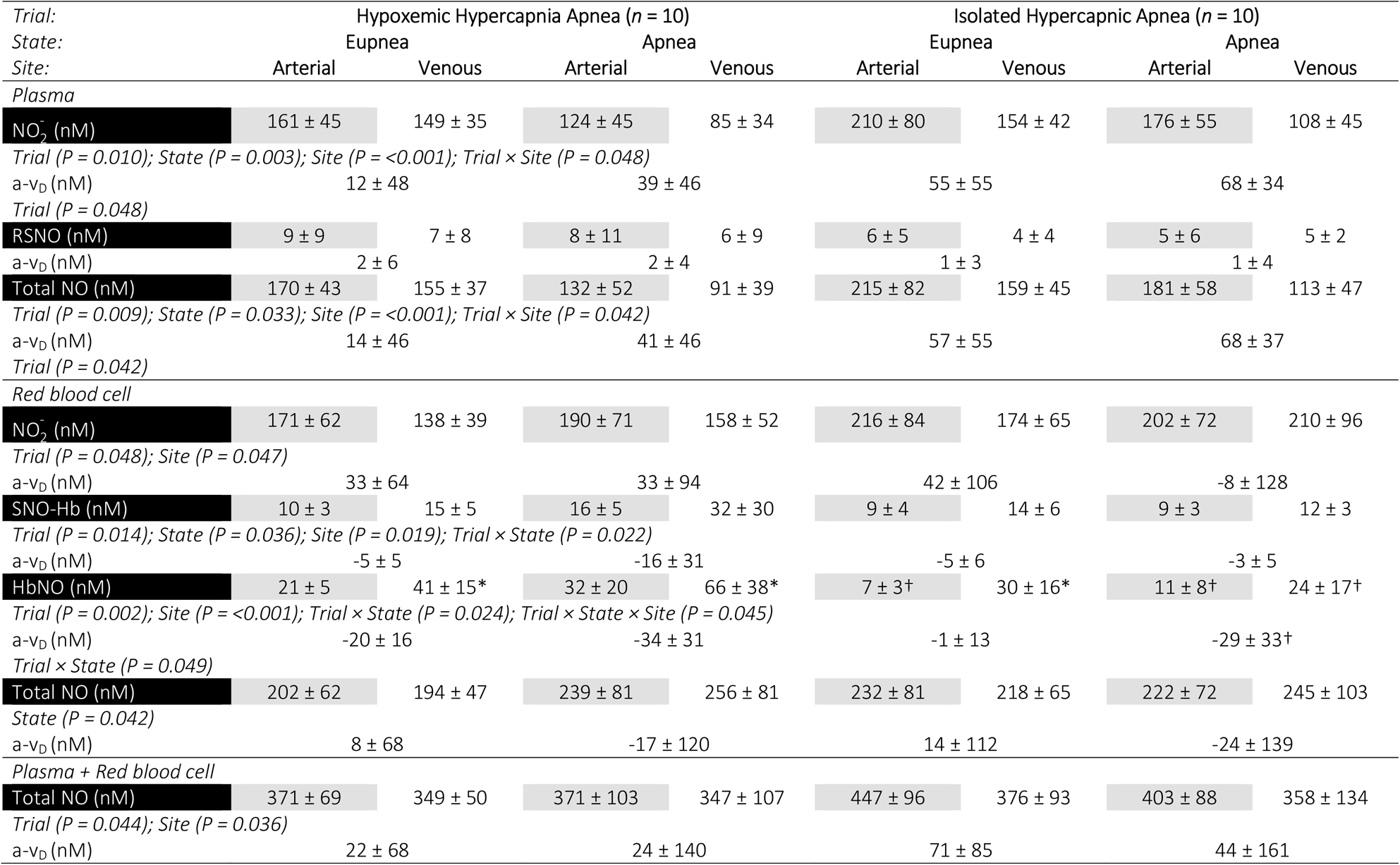
Nitrosative stress. Values are mean ± SD; a-v_D_,; NO, nitric oxide; 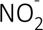, nitrite; RSNO, *S*-nitrosothiols; SNO-Hb, *S*-nitrosohemoglobin; HbNO, nitrosylhemoglobin. Positive or negative a-v_D_ reflects cerebral uptake (loss or consumption) or output (gain or formation) respectively. *different between site for given trial and state (*P* < 0.05); †different between trial for given state and site (*P* < 0.05).

| Trial:<br>State:<br>Site: | Hypoxemic Hypercapnia Apnea (n = 10) |  |  |  | Isolated Hypercapnic Apnea (n = 10) |  |  |  |
| --- | --- | --- | --- | --- | --- | --- | --- | --- |
|  | Eupnea |  | Apnea |  | Eupnea |  | Apnea |  |
|  | Arterial | Venous | Arterial | Venous | Arterial | Venous | Arterial | Venous |
| *Plasma* |  |  |  |  |  |  |  |  |
| **NO<sub>2</sub><sup>-</sup> (nM)** | 161 ± 45 | 149 ± 35 | 124 ± 45 | 85 ± 34 | 210 ± 80 | 154 ± 42 | 176 ± 55 | 108 ± 45 |
| *Trial (P = 0.010); State (P = 0.003); Site (P = <0.001); Trial × Site (P = 0.048)* |  |  |  |  |  |  |  |  |
| a-v<sub>D</sub> (nM) | 12 ± 48 |  | 39 ± 46 |  | 55 ± 55 |  | 68 ± 34 |  |
| *Trial (P = 0.048)* |  |  |  |  |  |  |  |  |
| **RSNO (nM)** | 9 ± 9 | 7 ± 8 | 8 ± 11 | 6 ± 9 | 6 ± 5 | 4 ± 4 | 5 ± 6 | 5 ± 2 |
| a-v<sub>D</sub> (nM) | 2 ± 6 |  | 2 ± 4 |  | 1 ± 3 |  | 1 ± 4 |  |
| **Total NO (nM)** | 170 ± 43 | 155 ± 37 | 132 ± 52 | 91 ± 39 | 215 ± 82 | 159 ± 45 | 181 ± 58 | 113 ± 47 |
| *Trial (P = 0.009); State (P = 0.033); Site (P = <0.001); Trial × Site (P = 0.042)* |  |  |  |  |  |  |  |  |
| a-v<sub>D</sub> (nM) | 14 ± 46 |  | 41 ± 46 |  | 57 ± 55 |  | 68 ± 37 |  |
| *Trial (P = 0.042)* |  |  |  |  |  |  |  |  |
| *Red blood cell* |  |  |  |  |  |  |  |  |
| **NO<sub>2</sub><sup>-</sup> (nM)** | 171 ± 62 | 138 ± 39 | 190 ± 71 | 158 ± 52 | 216 ± 84 | 174 ± 65 | 202 ± 72 | 210 ± 96 |
| *Trial (P = 0.048); Site (P = 0.047)* |  |  |  |  |  |  |  |  |
| a-v<sub>D</sub> (nM) | 33 ± 64 |  | 33 ± 94 |  | 42 ± 106 |  | -8 ± 128 |  |
| **SNO-Hb (nM)** | 10 ± 3 | 15 ± 5 | 16 ± 5 | 32 ± 30 | 9 ± 4 | 14 ± 6 | 9 ± 3 | 12 ± 3 |
| *Trial (P = 0.014); State (P = 0.036); Site (P = 0.019); Trial × State (P = 0.022)* |  |  |  |  |  |  |  |  |
| a-v<sub>D</sub> (nM) | -5 ± 5 |  | -16 ± 31 |  | -5 ± 6 |  | -3 ± 5 |  |
| **HbNO (nM)** | 21 ± 5 | 41 ± 15* | 32 ± 20 | 66 ± 38* | 7 ± 3† | 30 ± 16* | 11 ± 8† | 24 ± 17† |
| *Trial (P = 0.002); Site (P = <0.001); Trial × State (P = 0.024); Trial × State × Site (P = 0.045)* |  |  |  |  |  |  |  |  |
| a-v<sub>D</sub> (nM) | -20 ± 16 |  | -34 ± 31 |  | -1 ± 13 |  | -29 ± 33† |  |
| *Trial × State (P = 0.049)* |  |  |  |  |  |  |  |  |
| **Total NO (nM)** | 202 ± 62 | 194 ± 47 | 239 ± 81 | 256 ± 81 | 232 ± 81 | 218 ± 65 | 222 ± 72 | 245 ± 103 |
| *State (P = 0.042)* |  |  |  |  |  |  |  |  |
| a-v<sub>D</sub> (nM) | 8 ± 68 |  | -17 ± 120 |  | 14 ± 112 |  | -24 ± 139 |  |
| *Plasma + Red blood cell* |  |  |  |  |  |  |  |  |
| **Total NO (nM)** | 371 ± 69 | 349 ± 50 | 371 ± 103 | 347 ± 107 | 447 ± 96 | 376 ± 93 | 403 ± 88 | 358 ± 134 |
| *Trial (P = 0.044); Site (P = 0.036)* |  |  |  |  |  |  |  |  |
| a-v<sub>D</sub> (nM) | 22 ± 68 |  | 24 ± 140 |  | 71 ± 85 |  | 44 ± 161 |  |

## DISCUSSION

Local sampling of blood across the cerebral circulation during maximal apnea in world-class BH divers has provided important insight into the mechanisms underlying the redox-regulation of cerebral bioenergetic function during the most severe extremes of O_2_ demand/CO_2_ production recorded to date in unanesthetized and otherwise healthy humans^22^. Direct measures of intravascular OXNOS and inclusion of the IHA trial designed to prevent hypoxemia and better distinguish between competing vasoactive stimuli, highlights three novel findings. First, the combined elevation in the net cerebral output of A^·-^ and LOOH during apnea, collectively taken to reflect increased free radical-mediated lipid peroxidation, was more pronounced in HHA, highlighting a key contribution for hypoxemia. Second, this coincided with a lower apnea-induced elevation in gCBF and related suppression in plasma 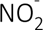 uptake, implying reduced consumption and delivery of NO consistent with increased OXNOS. Finally, that apnea-induced 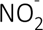 uptake and HbNO output prevailed in the face of increased SNO-Hb output and (general) RSNO uptake, tentatively suggests 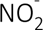 reduction as the more likely mechanism underlying endocrine NO vasoregulation with the capacity to transduce physiological O_2_-CO_2_ gradients into graded vasodilation. Collectively, these findings highlight the dynamic interplay that takes place between free radicals and NO metabolites during a single arteriovenous transit across the cerebral circulation and corresponding implications for cerebral bioenergetic function that may have clinical translational relevance.

### Hypoxemia is a key stimulus for cerebral OXNOS

We recently documented molecular evidence for structural destabilization of the NVU that was selectively more pronounced during apnea in HHA compared to IHA despite comparatively suppressed, albeit concerted vasodilation^6^. While this was originally attributed purely to the ‘mechanical’ stress imposed by more severe systemic and intracranial hypertension caused by hypoxemia, the present findings suggest a potentially additive role for ‘chemical’ stress, taking the form of elevated OXNOS, to which the human brain is especially vulnerable.

To do this, we employed EPR spectroscopic detection of the distinctive A^·-^ ‘doublet’ as a direct biomarker of global free radical formation with LOOH serving as a distal reactant of lipid peroxidation^9^. That net cerebral uptake (a>v) of ascorbate, α-tocopherol and (total) LSA was observed during apnea in both trials, and more pronounced in HHA, was taken to reflect enhanced sacrificial consumption (uptake by brain parenchyma and/or blood-borne reactions during circulatory transit) to constrain chain initiation-propagation reactions. However, this failed to prevent the overall shift in redox imbalance favouring increased free radical-mediated lipid peroxidation confirmed by proportional increases in the net cerebral output of A^·-^ and LOOH (v>a) that were selectively elevated in HHA. The greater elevation in A^·-^ output in HHA exceeded that predicted by prior modelling due to more ascorbate available for oxidation^23^ (given increased consumption), highlighting authentic (i.e. *de novo*) free radical formation. Our measured A^·-^ gradients in HHA (v = 37 % > a) also exceeded those previously recorded during HHA in a separate group of elite apneists^24^ (v = 4 % > a) and were even more pronounced than those documented during vigorous cycling exercise^9^ (v = 12 % > a) - helping place its magnitude into clearer physiological context.

While it is conceivable that hypoxemia, hypercapnia and hyperoxemia independently contributed towards global oxidative stress, it was not experimentally possible to fully differentiate between stimuli, only combinations thereof. Hypoxemia and hyperoxemia independently generate mitochondrial superoxide (O_2_^·-^) and NO^·^, notwithstanding contributions from extra-mitochondrial sources including nicotinamide adenine dinucleotide phosphate- and xanthine oxidases, cytochrome P-450 enzymes, lipoxygenases and phagocytes, that can further propagate oxidative stress via secondary formation of hydrogen peroxide (H_2_O_2_), peroxynitrite (ONOO^-^) and Haber-Weiss/Fenton catalyzed hydroxyl radicals (OH^·-^)^25, 26^. Furthermore, the addition of (hypercapnic) acidosis, common to both trials yet more severe in HX given the extended BH times, would have been expected to further amplify oxidation since molecular CO_2_ can react with ONOO^-^ and H_2_O_2_ to form redox reactive carbonate radicals and peroxymonocarbonate^27^. However, that cerebral oxidative stress was considerably more pronounced in HHA suggests a key catalytic role for hypoxemia that far outweighed the additive effects of hyperoxemia and (more severe) hypercapnia, confirming prior observations in healthy humans albeit exposed to poikilocapnic (hypocapnic) hypoxia^9, 28^.

To what extent this represents a physiologically adaptive (molecular signalling) or pathologically maladaptive (structurally damaging) response remains unclear given the pleiotropic mechanisms underlying redox-regulation of NVU function^4^. That oxidative stress was elevated in the face of reduced gCMRO_2_ and gCMR_Glucose_, attributable in part to hypercapnia^8^, has also been observed in (more) anoxia–hypoxia-tolerant vertebrates and interpreted to reflect an adaptive O_2_-conserving strategy to improve neuronal survival^29^. However, two observations in the present study tentatively suggest that the overall impact was likely maladaptive. First, that apnea in NX was associated with a lower net uptake of plasma 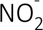 implied reduced consumption of NO, potentially tempering vasodilation (see below). This may reflect oxidative inactivation of NO by O ^·-^ and/or secondary lipid-derived alkoxyl-alkyl radicals formed subsequent to the reductive decomposition of (elevated) LOOH to form ONOO, collectively taken as evidence for elevated OXNOS (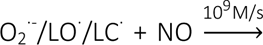 ONOO^-^)^30^. Second, that our prior research has shown that apnea induces mild disruption of the BBB and increased neuronal-gliovascular reactivity that were consistently more pronounced in HHA^6^, stands testament to the thermodynamic potential of radicals to cause structural damage to the NVU, which is not unreasonable given its histological vulnerability^4, 31^.

### NO metabolites and vasoregulation

Our findings highlight that the NO metabolite pool is in a perpetual state of dynamic flux even within the time constraints of a single a-v transit and that metabolite re-apportionment between plasma and RBC compartments is not solely related to changes in O_2_ but also involves CO_2_ and pH (Figure 3). By measuring corresponding gradients of 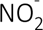 and SNO-Hb, the principal metabolites capable of transducing NO bioactivity within the cerebral microcirculation^32, 33^, we sought to determine if differential changes could further inform the mechanism(s) underlying cerebral vasodilation across extremes of PaO_2_ and PaCO_2_. Both metabolites likely originated from the combined effects of apnea-induced elevations in shear-mediated conduit vessel endothelial^34^, neuronal^35^ NO synthase bioactivation and transpulmonary *S*-nitrosylation^32^; however, their respective roles in endocrine NO signalling and cerebral bioenergetic function remain widely contested, albeit unified by allosteric-coupled Hb deoxygenation and acidic disproportionation^5^.

The 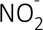 reductase hypothesis favors 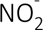 as the major intravascular NO storage molecule with conversion to NO or dinitrogen trioxide (N_2_O_3_) catalyzed by deoxyHb-mediated reduction and acidic disproportionation^13, 33, 36^. In contrast, the SNO-Hb hypothesis contends that NO is stabilized, transported, and delivered by intra-molecular NO group transfers between the heme iron and β-93 cysteine, that upon deoxygenation releases an as of yet unidentified moiety that is exported from the RBC involving transnitrosylation of the anion exchanger AE1, as a low molecular weight RSNO to effect vasodilation^32, 37, 38^.

The direction and magnitude of gradients observed are generally more consistent with those predicted by the 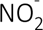 reductase (as opposed to the SNO-Hb) hypothesis. In support, systemic (a) plasma 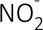 fell during apnea and was accompanied by a concomitant increase in net uptake implying consumption from artery to vein (a>v), although we cannot discount consistently elevated 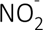 formation on the arterial side due to elevated NOS activity. Plasma 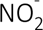 consumption was especially marked in HX, coinciding with the most pronounced elevation in gCBF and where the total NO metabolite pool was greatest. This was accompanied by NO formation with concomitant output (v>a) of HbNO, albeit suppressed in IHA consistent with the prevailing SO_2_ since hyperoxemia persisted at end apnea, culminating in an overall net gain in NO metabolites. These findings were taken to reflect a combination of deoxyHb-mediated reduction of 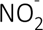 in the RBC and acidic disproportionation subsequent to marked hypercapnia-induced acidosis. Local formation of NO, potentially compounded by the observed increase in ascorbate consumption^39^, binds to vicinal deoxyHb yielding HbNO (Equations 1 and 2) and, to a lesser extent, the intermediate nitrosating species dinitrogen trioxide that can escape the RBC to effect graded vasodilation^33^:

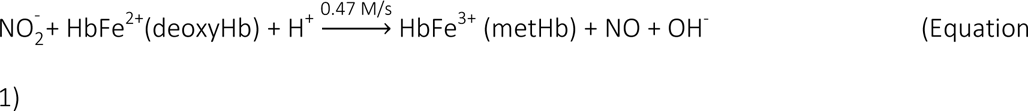

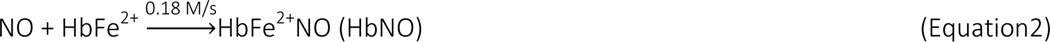

The gradients observed reproduce those previously documented across the exercising human forearm circulation and 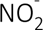 that originally informed the 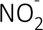 reductase hypothesis^33, 36^. Similar gradients were more recently documented across the cerebral circulation at rest and in response to acute exercise during hypocapnic hypoxia^13^.

In contrast, that apnea stimulated SNO-Hb output (v>a) and RSNO uptake (a>v) argues against the SNO-Hb hypothesis, since the gradients observed were diametrically opposed to those originally predicted^40^, yet consistent with the observations of other research laboratories^41, 42^. We would have expected an allosterically mediated release from a-v during circulatory transit, resulting in a drop in the RBC and corresponding rise in plasma RSNO in the venous circulation, consistent with SNO-Hb consumption (a>v) and RSNO delivery (v>a) during local deoxygenation. Yet, the gradients observed consistently reflected SNO-Hb formation (v>a) and RSNO consumption (a>v), that in the former, was further compounded during apnea and unrelated to the increase in gCBF.

### Experimental limitations

Several limitations warrant consideration. First, it was not possible to fully disassociate changes in PaO_2_ from PaCO_2_, only combinations thereof, given the inability to ‘match’ end-apnea hypercapnia that was unavoidably more pronounced in HX owing to extended BH times. Second, it was unfortunate that we did not measure plasma 3-nitrotyrosine due to inadequate sample volume: its elevation would have indirectly confirmed increased ONOO^-^ formation subsequent to oxidative inactivation of NO underlying OXNOS, consistent with our prior observations in hypoxia^35^. Third, the profiling of NO metabolite gradients, while informative, fails to unequivocally identify the specific NO species responsible for conserving bioactivity and the mechanism(s) underpinning vasoregulation, including potential contributions from ATP-mediated activation of P_2Y_ receptors. Finally, despite prospective power calculations informed by our prior research^24^, we acknowledge the small sample sizes employed may have limited our ability to detect treatment effects, albeit challenging to perform large-scale invasive studies of this nature. Caveats notwithstanding, mapping the dynamic transvascular interplay of OXINOS biomarkers in this applied model of human asphyxia may prove clinically relevant in that it may help define more sensitive biomarkers of cerebrovascular health and inform interventions designed to optimize tissue oxygenation in the critically ill.

## Data Availability

The data that support the findings of this study are not openly available due to reasons of sensitivity and are available from the corresponding author upon reasonable request.

## SUPPLEMENTAL MATERIAL

**Supplemental Table 1.**
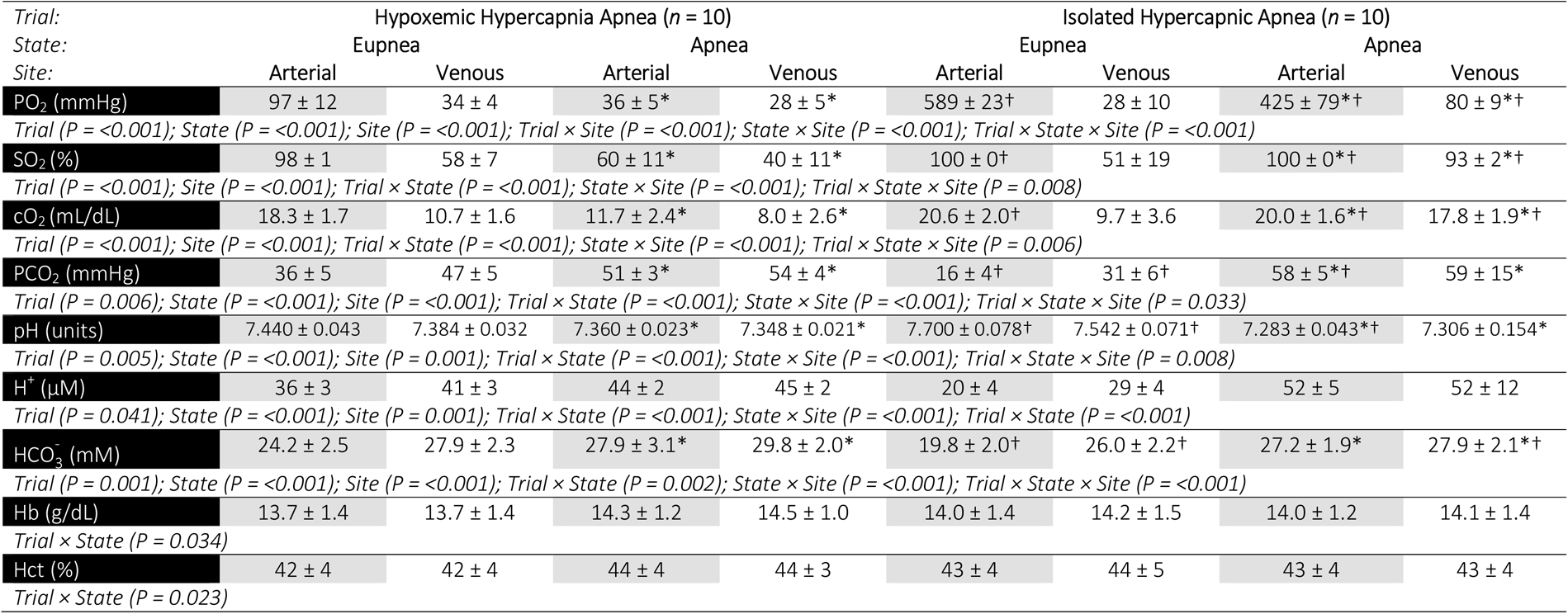
Blood gas variables. Values are mean ± SD; PO_2_/PCO_2_, partial pressure of oxygen/carbon dioxide; SO_2_, oxyhemoglobin saturation; cO, oxygen content; H^+^, hydrogen ions 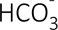, bicarbonate ions; Hb, hemoglobin; Hct, hematocrit. *different between state for given trial and site (*P* < 0.05); †different between trial for given state and site (*P* < 0.05).

**Supplemental Table 2.** Cardiopulmonary function. Values are mean ± SD; HR, heart rate; SV, stroke volume; 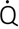, cardiac output; MAP, mean arterial pressure; IJVP, internal jugular venous pressure; CPP, cerebral perfusion pressure. Δ, apnea minus eupnea; *different between state for given trial (*P* < 0.05); †different between trial for given state or Δ (*P* < 0.05).

| *Trial:* | Hypoxemic Hypercapnia Apnea (*n* = 10) |  |  | Isolated Hypercapnic Apnea (*n* = 10) |  |  |
| --- | --- | --- | --- | --- | --- | --- |
| *State:* | Eupnea | Apnea | Δ | Eupnea | Apnea | Δ |
| **HR (b/min)** | 64 ± 7 | 59 ± 14 | -5 ± 12 | 65 ± 11 | 69 ± 13 | 4 ± 18 |
| **SV (mL/min)** | 78 ± 16 | 67 ± 16* | -11 ± 15 | 86 ± 17† | 90 ± 17† | 5 ± 12† |
| *Trial (P = 0.001); Trial × State (P = 0.001)* |  |  |  |  |  |  |
| **Q̇ (L/min)** | 4.92 ± 0.91 | 3.91 ± 1.17* | -1.01 ± 1.21 | 5.46 ± 1.01 | 6.29 ± 1.97† | 0.83 ± 1.73† |
| *Trial (P = 0.001); Trial × State (P = 0.029)* |  |  |  |  |  |  |
| **MAP (mmHg)** | 116 ± 8 | 177 ± 8* | 61 ± 9 | 111 ± 8† | 158 ± 16*† | 47 ± 14*† |
| *Trial (P = 0.001); State (P = <0.001); Trial × State (P = 0.018)* |  |  |  |  |  |  |
| **IJVP (mmHg)** | 10 ± 2 | 22 ± 1* | 12 ± 2 | 10 ± 2 | 17 ± 5*† | 7 ± 5*† |
| *Trial (P = 0.015); State (P = <0.001); Trial × State (P = 0.016)* |  |  |  |  |  |  |
| **CPP (mmHg)** | 106 ± 9 | 155 ± 7 | 49 ± 8 | 101 ± 9 | 141 ± 16 | 40 ± 13 |
| *Trial (P = 0.002); State (P = <0.001); Trial × State (P = 0.112)* |  |  |  |  |  |  |

**Supplemental Table 3.** Cerebral bioenergetic function. Values are mean ± SD; 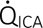, internal carotid artery flow; 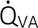, vertebral artery flow; gCBF, global cerebral blood flow; gCDO_2_, global cerebral delivery of oxygen; gCD_Glucose_, global cerebral delivery of glucose; CMR, cerebral metabolic rate; Δ, apnea minus eupnea. *different between state for given trial (*P* < 0.05); †different between trial for given state (*P* < 0.05).

| Trial: | Hypoxemic Hypercapnia Apnea (n = 10) |  |  | Isolated Hypercapnic Apnea (n = 10) |  |  |
| --- | --- | --- | --- | --- | --- | --- |
| State: | Eupnea | Apnea | Δ | Eupnea | Apnea | Δ |
| Perfusion |  |  |  |  |  |  |
| Q<sub>ICA</sub> (mL/min) | 458 ± 55 | 848 ± 148* | 390 ± 119 | 317 ± 92† | 938 ± 211* | 621 ± 174† |
| State (P = <0.001); Trial × State (P = 0.001) |  |  |  |  |  |  |
| Q<sub>VA</sub> (mL/min) | 177 ± 82 | 307 ± 133* | 130 ± 54 | 113 ± 55† | 357 ± 198* | 244 ± 146† |
| State (P = <0.001); Trial × State (P = 0.009) |  |  |  |  |  |  |
| gCBF (mL/min) | 635 ± 84 | 1155 ± 147* | 520 ± 114 | 31 ± 7† | 92 ± 22* | 865 ± 260† |
| State (P = <0.001); Trial × State (P = 0.002) |  |  |  |  |  |  |
| Metabolism |  |  |  |  |  |  |
| gCDO<sub>2</sub> (mL/min) | 116 ± 22 | 137 ± 43* | 20 ± 30 | 88 ± 20† | 259 ± 63*† | 171 ± 52† |
| Trial (P = <0.001); State (P = <0.001); Trial × State (P = <0.001) |  |  |  |  |  |  |
| gCD<sub>Gluc</sub> (mM/min) | 3.45 ± 0.60 | 6.30 ± 1.07* | 2.85 ± 0.77 | 2.20 ± 0.49† | 7.48 ± 1.57*† | 5.28 ± 1.40† |
| State (P = <0.001); Trial × State (P = <0.001) |  |  |  |  |  |  |
| gCMRO<sub>2</sub> (mL/min) | 47 ± 8 | 43 ± 12 | -5 ± 17 | 48 ± 19 | 28 ± 8 | -19 ± 18 |
| State (P = 0.015) |  |  |  |  |  |  |
| gCMR<sub>Glucose</sub> (mM/min) | 0.37 ± 0.09 | 0.30 ± 0.09 | -0.07 ± 0.14 | 0.47 ± 0.14 | 0.50 ± 0.52 | 0.04 ± 0.58 |
| Trial (P = 0.044) |  |  |  |  |  |  |

## ACKNOWLEDGMENTS

The authors acknowledge the cheerful cooperation of all participants. We are also indebted to Professor IS Young and Dr Jane McEneny for specialist technical input.

## REFERENCES

1. Schaeffer S and Iadecola C. Revisiting the neurovascular unit. Nat Neurosci. 2021;24:1198–1209.

2. Rinaldi C, Donato L, Alibrandi S, Scimone C, D’Angelo R and Sidoti A. Oxidative Stress and the Neurovascular Unit. Life (Basel). 2021;11.

3. Bailey DM. Oxygen, evolution and redox signalling in the human brain; quantum in the quotidian. The Journal of physiology. 2019;597:15–28.

4. Cobley JN, Fiorello ML and Bailey DM. 13 reasons why the brain is susceptible to oxidative stress. Redox biology. 2018;15:490–503.

5. Gladwin MT and Schechter AN. NO contest: nitrite versus S-nitroso-hemoglobin. Circulation Research. 2004;94:851–5.

6. Bailey DM, Bain AR, Hoiland RL, Barak OF, Drvis I, Hirtz C, Lehmann S, Marchi N, Janigro D, MacLeod DB, Ainslie PN and Dujic Z. Hypoxemia increases blood-brain barrier permeability during extreme apnea in humans. Journal of cerebral blood flow and metabolism : official journal of the International Society of Cerebral Blood Flow and Metabolism. 2022;42:1120–1135.

7. WMA. World Medical Association Declaration of Helsinki: ethical principles for medical research involving human subjects. Journal of the American Medical Association. 2013;310:2191–4.

8. Bain AR, Ainslie PN, Barak OF, Hoiland RL, Drvis I, Mijacika T, Bailey DM, Santoro A, DeMasi DK, Dujic Z and MacLeod DB. Hypercapnia is essential to reduce the cerebral oxidative metabolism during extreme apnea in humans. Journal of Cerebral Blood Flow and Metabolism. 2017:271678X16686093.

9. Bailey DM, Rasmussen P, Evans KA, Bohm AM, Zaar M, Nielsen HB, Brassard P, Nordsborg NB, Homann PH, Raven PB, McEneny J, Young IS, McCord JM and Secher NH. Hypoxia compounds exercise-induced free radical formation in humans; partitioning contributions from the cerebral and femoral circulation. Free Radical Biology & Medicine. 2018;124:104–113.

10. Vuilleumier JP and Keck E. Fluorimetric assay of vitamin C in biological materials using a centrifugal analyser with fluorescence attachment. Journal of Micronutritional Analysis. 1993;5:25–34.

11. Catignani GL and Bieri JG. Simultaneous determination of retinol and alpha-tocopherol in serum or plasma by liquid chromatography. Clinical Chemistry. 1983;29:708–712.

12. Thurnham DI, Smith E and Flora PS. Concurrent liquid-chromatographic assay of retinol, *a*-tocopherol, *β*-carotene, *a*-carotene, lycopene, and *β*-cryptoxanthin in plasma, with tocopherol acetate as internal standard. Clinical Chemistry. 1988;34:377–381.

13. Bailey DM, Rasmussen P, Overgaard M, Evans KA, Bohm AM, Seifert T, Brassard P, Zaar M, Nielsen HB, Raven PB and Secher NH. Nitrite and *S*-Nitrosohemoglobin exchange across the human cerebral and femoral circulation: relationship to basal and exercise blood flow responses to hypoxia. Circulation. 2017;135:166–176.

14. Rogers SC, Khalatbari A, Gapper PW, Frenneaux MP and James PE. Detection of human red blood cell-bound nitric oxide. Journal of Biological Chemistry. 2005;280:26720–26728.

15. Pinder AG, Rogers SC, Khalatbari A, Ingram TE and James PE. The measurement of nitric oxide and its metabolites in biological samples by ozone-based chemiluminescence. Methods in molecular biology. 2009;476:10–27.

16. Wesseling KH, Jansen JR, Settels JJ and Schreuder JJ. Computation of aortic flow from pressure in humans using a nonlinear, three-element model. Journal of applied physiology. 1993;74:2566–73.

17. Woodman RJ, Playford DA, Watts GF, Cheetham C, Reed C, Taylor RR, Puddey IB, Beilin LJ, Burke V, Mori TA and Green D. Improved analysis of brachial artery ultrasound using a novel edge-detection software system. Journal of applied physiology. 2001;91:929–37.

18. Willie CK, Macleod DB, Shaw AD, Smith KJ, Tzeng YC, Eves ND, Ikeda K, Graham J, Lewis NC, Day TA and Ainslie PN. Regional brain blood flow in man during acute changes in arterial blood gases. Journal of Physiology. 2012;590:3261–75.

19. Hoiland RL, Ainslie PN, Bain AR, MacLeod DB, Stembridge M, Drvis I, Madden D, Barak O, MacLeod DM and Dujic Z. Beta1-Blockade increases maximal apnea duration in elite breath-hold divers. Journal of applied physiology. 2017;122:899–906.

20. Willie CK, Ainslie PN, Drvis I, MacLeod DB, Bain AR, Madden D, Maslov PZ and Dujic Z. Regulation of brain blood flow and oxygen delivery in elite breath-hold divers. Journal of Cerebral Blood Flow and Metabolism. 2015;35:66–73.

21. Myerson A and Loman J. Internal jugular venous pressure in man: Its relationship to cerebrospinal fluid and carotid arterial pressures. Archives of Neurology and Psychiatry. 1932;27:836–846.

22. Bailey DM, Willie CK, Hoiland RL, Bain AR, MacLeod DB, Santoro MA, DeMasi DK, Andrijanic A, Mijacika T, Barak OF, Dujic Z and Ainslie PN. Surviving without oxygen: how low can the human brain go? High altitude medicine & biology. 2017;18:73–79.

23. Fall L, Brugniaux JV, Davis D, Marley CJ, Davies B, New KJ, McEneny J, Young IS and Bailey DM. Redox-regulation of haemostasis in hypoxic exercising humans: a randomised double-blind placebo-controlled antioxidant study. The Journal of physiology. 2018;596:4879–4891.

24. Bain AR, Ainslie PN, Hoiland RL, Barak OF, Drvis I, Stembridge M, MacLeod DM, McEneny J, Stacey BS, Tuaillon E, Marchi N, De Maudave AF, Dujic Z, MacLeod DB and Bailey DM. Competitive apnea and its effect on the human brain: focus on the redox regulation of blood-brain barrier permeability and neuronal-parenchymal integrity. FASEB journal : official publication of the Federation of American Societies for Experimental Biology. 2018;32:2305–2314.

25. Turrens JF. Mitochondrial formation of reactive oxygen species. Journal of Physiology. 2003;552.2:335–344.

26. Murphy MP. How mitochondria produce reactive oxygen species. The Biochemical journal. 2009;417:1–13.

27. Augusto O and Truzzi DR. Carbon dioxide redox metabolites in oxidative eustress and oxidative distress. Biophys Rev. 2021;13:889–891.

28. Bailey DM, Taudorf S, Berg RMG, Lundby C, McEneny J, Young IS, Evans KA, James PE, Shore A, Hullin DA, McCord JM, Pedersen BK and Moller K. Increased cerebral output of free radicals during hypoxia: implications for acute mountain sickness? American Journal of Physiology (Regulatory, Integrative and Comparative Physiology). 2009;297:R1283–1292.

29. Bailey DM. Oxygen and brain death; back from the brink. Experimental physiology. 2019;104:1769–1779.

30. Bailey DM, Young IS, McEneny J, Lawrenson L, Kim J, Barden J and Richardson RS. Regulation of free radical outflow from an isolated muscle bed in exercising humans. American Journal of Physiology (Heart and Circulatory Physiology). 2004;287:H1689–H1699.

31. Janigro D, Bailey DM, Lehmann S, Badaut J, O’Flynn R, Hirtz C and Marchi N. Peripheral blood and salivary biomarkers of blood–brain barrier permeability and neuronal damage: clinical and applied concepts. Frontiers in Neurology. 2021.

32. Jia L, Bonaventura C, Bonaventura J and Stamler JS. S-nitrosohaemoglobin: a dynamic activity of blood involved in vascular control. Nature. 1996;380:221–6.

33. Cosby K, Partovi KS, Crawford JH, Patel RP, Reiter CD, Martyr S, Yang BK, Waclawiw MA, Zalos G, Xu X, Huang KT, Shields H, Kim-Shapiro DB, Schechter AN, Cannon RO and Gladwin MT. Nitrite reduction to nitric oxide by deoxyhemoglobin vasodilates the human circulation. Nature Medicine. 2003;9:1498–505.

34. Dimmeler S, Fleming I, Fisslthaler B, Hermann C, Busse R and Zeiher AM. Activation of nitric oxide synthase in endothelial cells by Akt-dependent phosphorylation. Nature. 1999;399:601–5.

35. O’Gallagher K, Rosentreter RE, Elaine Soriano J, Roomi A, Saleem S, Lam T, Roy R, Gordon GR, Raj SR, Chowienczyk PJ, Shah AM and Phillips AA. The Effect of a Neuronal Nitric Oxide Synthase Inhibitor on Neurovascular Regulation in Humans. Circ Res. 2022;131:952–961.

36. Gladwin MT, Shelhamer JH, Schechter AN, Pease-Fye ME, Waclawiw MA, Panza JA, Ognibene FP and Cannon RO, III. Role of circulating nitrite and S-nitrosohemoglobin in the regulation of regional blood flow in humans. Proceedings of the National Academy of Sciences of the United States of America. 2000;97:11482–11487.

37. Stamler J, Jia L, Eu J, McMahon T, Demchenko I, Bonaventura J, Gernet K and Piantadosi C. Blood Flow Regulation by *S*-Nitrosohemoglobin in the Physiological Oxygen Gradient. Science. 1997;276:2034–2037.

38. Pawloski JR, Hess DT and Stamler JS. Export by red blood cells of nitric oxide bioactivity. Nature. 2001;409:622–6.

39. Ladurner A, Schmitt CA, Schachner D, Atanasov AG, Werner ER, Dirsch VM and Heiss EH. Ascorbate stimulates endothelial nitric oxide synthase enzyme activity by rapid modulation of its phosphorylation status. Free radical biology & medicine. 2012;52:2082–90.

40. McMahon TJ, Moon RE, Luschinger BP, Carraway MS, Stone AE, Stolp BW, Gow AJ, Pawloski JR, Watke P, Singel DJ, Piantadosi CA and Stamler JS. Nitric oxide in the human respiratory cycle. Nature Medicine. 2002;8:711–7.

41. Gladwin MT, Wang X, Reiter CD, Yang BK, Vivas EX, Bonaventura C and Schechter AN. S-Nitrosohemoglobin is unstable in the reductive erythrocyte environment and lacks O2/NO-linked allosteric function. The Journal of biological chemistry. 2002;277:27818–28.

42. Wang X, Bryan NS, MacArthur PH, Rodriguez J, Gladwin MT and Feelisch M. Measurement of nitric oxide levels in the red cell: validation of tri-iodide-based chemiluminescence with acid-sulfanilamide pretreatment. The Journal of biological chemistry. 2006;281:26994–7002.

43. Huang Z, Shiva S, Kim-Shapiro DB, Patel RP, Ringwood LA, Irby CE, Huang KT, Ho C, Hogg N, Schechter AN and Gladwin MT. Enzymatic function of hemoglobin as a nitrite reductase that produces NO under allosteric control. Journal of Clinical Investigation. 2005;115:2099–107.

44. Guzy R, Hoyos B, Robin E, Chen H, Liu L, Mansfield K, Simon M, Hammerling U and Schumacker P. Mitochondrial complex III is required for hypoxia-induced ROS production and cellular oxygen sensing. Cell Metabolism. 2005;1:401–8.

45. Williams NH and Yandell JK. Outer-sphere electron-transfer reactions of ascorbic anions. Australian Journal of Chemistry. 1982;35:1133–1144.

46. Buettner GR. The pecking order of free radicals and antioxidants: lipid peroxidation, a-tocopherol, and ascorbate. Archives of Biochemistry and Biophysics. 1993;300:535–543.

